# Identification of Spared and Proportionally Controllable Hand Motor Dimensions in Motor Complete Spinal Cord Injuries Using Latent Manifold Analysis

**DOI:** 10.1101/2024.05.28.24307964

**Authors:** Raul C. Sîmpetru, Daniela Souza de Oliveira, Matthias Ponfick, Alessandro Del Vecchio

**Affiliations:** Department Artificial Intelligence in Biomedical Engineering, Friedrich-Alexander-Universität Erlangen-Nürnberg, 91052 Erlangen, Germany; Querschnittzentrum Rummelsberg, Krankenhaus Rummelsberg GmbH, 90592 Schwarzenbruck, Germany

**Keywords:** SCI, Rehabilitation, EMG, AI

## Abstract

The loss of bilateral hand function is a debilitating challenge for millions of individuals that suffered a motor-complete spinal cord injury (SCI). We have recently demonstrated in eight tetraplegic individuals the presence of highly functional spared spinal motor neurons in the extrinsic muscles of the hand that are still capable of generating proportional flexion and extension signals. In this work, we hypothesized that an artificial intelligence (AI) system could automatically learn the spared electromyographic (EMG) patterns that encode the attempted movements of the paralyzed digits. We constrained the AI to continuously output the attempted movements in the form of a digital hand so that this signal could be used to control any assistive system (e.g., exoskeletons, electrical stimulation). We trained a convolutional neural network using data from 13 uninjured (control) participants and 8 motor-complete tetraplegic participants to study the latent space learned by the AI. Our model can automatically differentiate between eight different hand movements, including individual finger flexions, grasps, and pinches, achieving a mean accuracy of 98.3% within the SCI group. Moreover, the model could distinguish with 100% accuracy whether a participant had an injury or not, and it could also facilitate proportional control of certain movements after the injury. Analysis of the latent space of the model revealed that proportionally controllable movements exhibited an elliptical path, while movements lacking proportional control followed a chaotic trajectory. We found that proportional control of a movement can only be correctly estimated if the latent space embedding of the movement follows an elliptical path (correlation = 0.73; p *<* 0.001). These findings emphasize the reliability of the proposed system for closed-loop applications that require an accurate estimate of the spinal cord motor output.

## 1 Introduction

LOSING hand motor functions is the tragic reality of 10.85 million (133 out of 100,000) people with cervical spinal cord injury (SCI) worldwide [1]. In the spinal cord, *α*-motoneurons serve as connections between the central and peripheral nervous system with a diverse array of skeletal muscles. Motor-complete SCI is typically characterized by profound paralysis and a lack of discernible volitional control in all limbs below the area of injury [2]. Recently, we have used surface electromyography (sEMG), a noninvasive neural interface [3], to investigate muscle activity below the injury level in individuals who have been clinically labeled as having a cervical motor-complete SCI [2]. We found that these individuals can precisely control the activity of several spared motor units (MU) during intentionally attempted hand digit movements [4], [5]. Although the presence of EMG activity in individuals with motor-complete SCI was demonstrated already several years ago [6], only recently were we able to demonstrate that this activity can be controlled in real-time and at the level of individual MUs. We showed that eight individuals with chronic SCI could proportionally modulate the activity of motor neurons that control the movements of paralyzed digits [5].

Previous research has effectively classified various movements among individuals with SCI who maintained partial control of the hand [7], [8]. However, for those with motor-complete SCI, efforts have focused mainly on broader arm movements such as elbow flexion and extension [9]. Despite these advances, the precise classification or continuous prediction of intricate finger movements that are crucial for daily tasks remains a problem to be solved [10].

In this work (Fig. 1), we hypothesize that an artificial intelligence (AI) system could automatically map and decode individual finger movements from spared EMG signals of 8 motor-complete participants with SCI. The potential of having an AI system that automatically learns the EMG activation patterns generating specific movements has numerous applications for the recovery of the hand function, considering the large number of motor dimensions embedded in the hand. For this, we adapted our previously published deep learning model capable of realtime, accurate and proportional hand kinematics prediction for healthy participants [11] and tested it on 13 uninjured (control group) and 8 individuals suffering from a motor-complete SCI (SCI group).

**Fig. 1:**
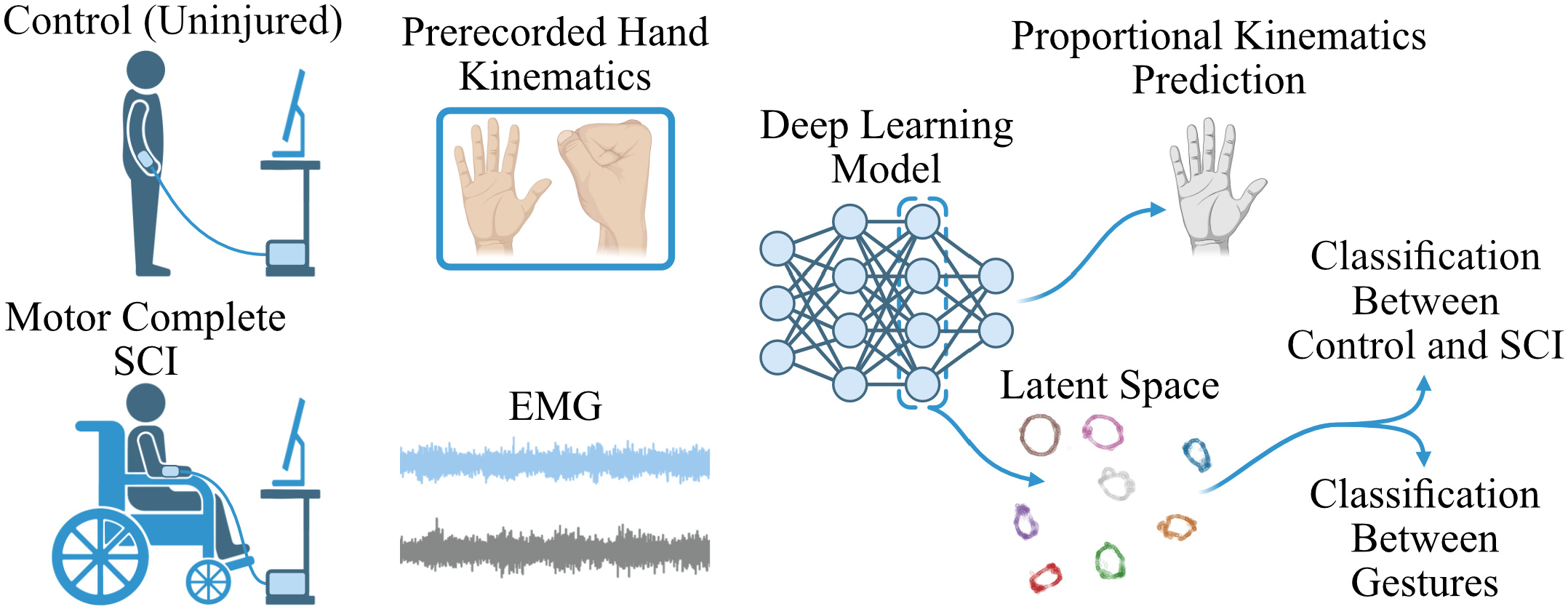
We collected surface electromyography (sEMG) data from a group of 8 individuals with cervical motor-complete spinal cord injuries (SCI) and 13 uninjured individuals (control). During the data collection process, participants were asked to attempt pre-recorded hand movements displayed on a screen. Using the data collected, we trained participant-specific deep learning models for the proportional prediction of the hand positions in a 3D space. Successively, we applied a dimensionality reduction technique called UMAP to transform the latent space into a 2D representation. The 2D projections were then used to distinguish between different hand movements for the same participant and whether the participant had an SCI or not. This analysis has the potential to offer people living with SCI the ability to control and perform multiple different movements with assistive systems. Moreover, the analysis of the latent space of the model revealed a preserved neural representation of the movement of the hand digits between healthy and injured participants.

We demonstrate that our system can proportionally predict all attempted movements in the control group and that some hand movements can also be proportionally predicted in individuals with motor-complete SCI. Furthermore, our objective was to understand the control signals generated by the neural network. We projected the high-dimensional latent space of the neural network using UMAP [12], a dimensionality reduction technique that preserves the intrinsic manifold of the data, into human-readable 2D. Using 2D projections, we were able to classify the 8 attempted movements with an accuracy greater than 93% in participants with SCI, and with 100% accuracy in participants without SCI only based on EMG signals. Comparison of the latent space between the control and SCI groups also highlights the potential of some movements to be fully regained by spinal cord injured participants (Fig. 1). This is of particular interest because, although there are spared MUs after injury, it is not known if there are sufficient agonist and antagonist MUs that allow proportional control. Interestingly, the AI model mapped proportionality with circularity. Movements that were proportionally predicted were mapped with a continuous circular trajectory for both the control and the SCI group. For some movements of the SCI group, the trajectories appeared to be chaotic, probably due to these movements lacking sufficient neural activity for proportional control. We show that the deviation error of a circular path in the projection of the latent space is negatively and significantly correlated with the accuracy of the proportional prediction (-0.73, p *<* 0.001). The proposed method could provide numerous advantages in clinical settings. For example, by analyzing the projection of the latent space of the neural network, it is possible to characterize how many motor dimensions are present in the EMG signal after the injury.

## 2 Methods

### 2.1 Datasets

Two datasets were used to investigate the potential use of the proposed AI system to automatically detect the spared motor dimensions that are still present in the high-density EMG of participants with SCI.

The first dataset served as the control group and consists of 13 young, uninjured adult participants (age 25.9 ± 2.8 years) (data also used in [13]– 15]).

The test group consists of 8 individuals with motor-complete SCI (the same dataset was also used in [5]). Table 1 shows the individual patient characteristics; however, in summary, all injuries occurred in the cervical area (C5-C6), 4 subjects have an AIS [2] grade of B (S1-3, 8), 3 of A (S4-6), and 1 of C (S7).

**TABLE 1:** Characteristics of the spinal cord injured participants. AIS = ASIA Injury Scale [2], Time Since Injury = The amount of time a subject has lived with an SCI at the time of recording, Sensory Level = The lowest spinal cord section that still has normal sensory function, MAS = Modified Ashworth Scale; R = Right, L = Left; spasticity was assessed for elbow flexion. More details in Souza de Oliveira *et al*. [5].

| Subject | Age range (years) | Gender | Injury Level | AIS | Time Since Injury (years) | Sensory Level | Spasticity in Upper Limb (MAS) |
| --- | --- | --- | --- | --- | --- | --- | --- |
| 1 | 36–40 | M | C6 | B | 18.8 | S5 | 0 |
| 2 | 31–35 | M | C5 | B | 9.1 | C5 | 0 |
| 3 | 41–45 | F | C6 | B | 24.2 | C6 | 0 |
| 4 | 36–40 | F | C5 | A | 24.2 | C5 | 0 |
| 5 | 31–35 | M | C6 | A | 22.2 | C6 | 0 |
| 6 | 56–60 | M | C5 | A | 6.9 | T3 | R: 2, L: 0 |
| 7 | 41–45 | M | C6 | C | 18.2 | C6 | R: 2, L: 0 |
| 8 | 36–40 | F | C5 | B | 5.0 | T1 | 1 |

All subjects were instructed to perform a series of grasps and individual digit movements, starting and ending each cycle in a specified position, or, in the case of individuals with motor-complete SCI, to attempt these movements. The starting position is defined as the natural resting position of the hand, and the end position is defined as the fully flexed position. The series consisted of individual digits flexion and extension, grasping, and two- and three-finger pinches (Fig. 2A). These movements were displayed in front of the subjects in the form of a virtual hand performing prerecorded movements at a frequency of 0.5 Hz (Fig. 2A & B). The virtual hand was a digital rendering of one representative hand from an uninjured participant. During movement, monopolar EMG signals were recorded from the forearm of the subjects at 2048 Hz. For the recordings we used three 8 by 8 electrode grids with 10 mm interelectrode distance (IED) (GR10MM0808, OT Bioelettronica, Turin, Italy) and two 13 by 5 grids with 8 mm IED (GR08MM1305, OT Bioelettronica, Turin, Italy), for a total of 320 electrodes. The 8 by 8 grids were applied around the upper forearm area below the elbow, while the 13 by 5 grids were placed above the wrist around the arm covering the dorsal and the ventral part of the forearm (Fig. 2B).

**Fig. 2:**
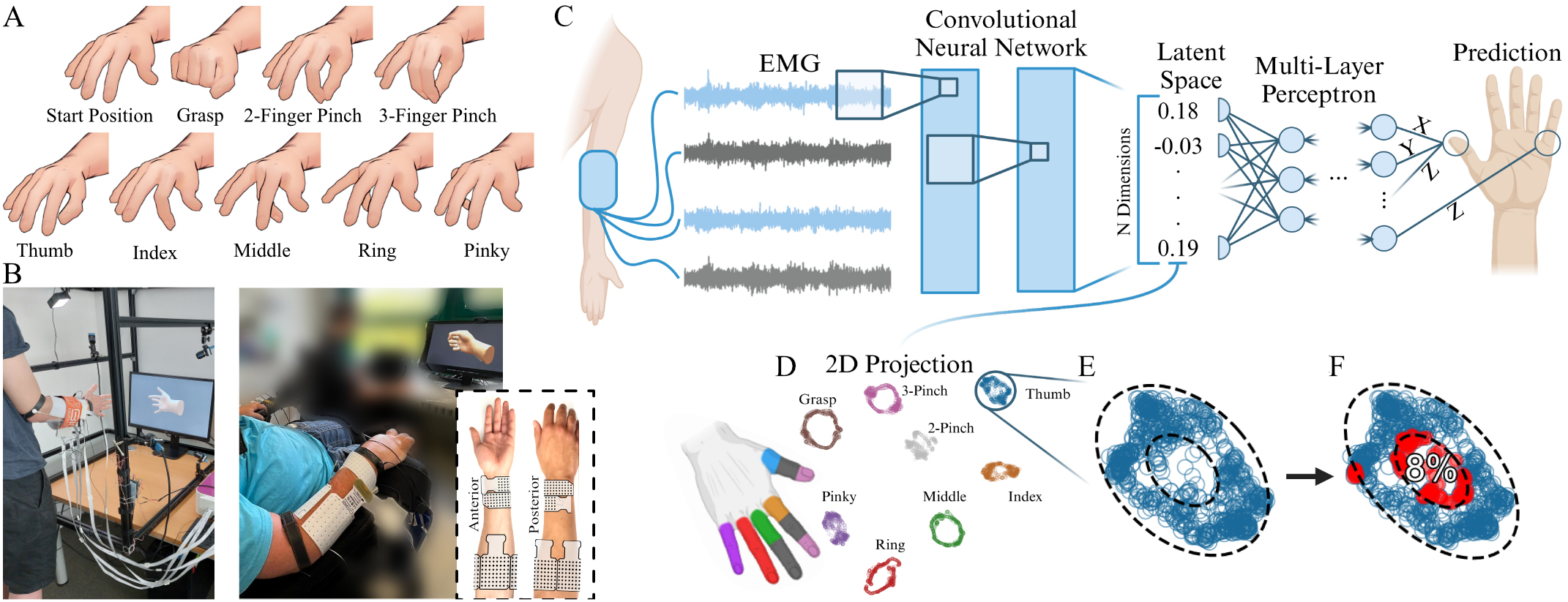
**A**. The various hand tasks performed in this study by the control (thirteen healthy uninjured participants) and eight participants with motor-complete SCI. The movements were executed at 0.5 Hz. **B**. Experimental setup is shown on both an uninjured and an injured participant. We used five electrode grids. Three 8 by 8 grids were placed on the proximal part of the forearm and two 13 by 5s above the wrist. **C**. Schematic overview of the deep learning model. Convolutional layers scan EMG signals in spatial and temporal dimension, detecting motor unit and muscle fiber action potential ensembles indicative of specific movement phases. Subsequent layers compress and integrate these overlaps, considering both intragrid and inter-grid information. The output of the convolutional layers is a high-dimensional latent space that separates and preserves the information of each movement. For each 100 ms EMG window, the convolutional network produces an n-dimensional vector that contains compressed information about the movement. The last part of the model is a multilayer perceptron that maps the high-dimensional latent space into the desired 3D coordinates. A detailed explanation can be found in Sîmpetru *et al*. [11]. **D**. Using UMAP, a non-linear dimensionality reduction technique that preserves the local and global information, we project the latent space vectors into human understandable 2D. The manifolds are color-coded according to the individual movements. For the projection metric, we computed the cosine distance to cluster similar vectors irrespective of their amplitude. Interestingly, the movement clusters are positioned in the same way as the fingers of the human hand. In addition, the grasping action is enclosed by the flexion and extension of all individual fingers. **E**. For each manifold, we fit an inner and outer ellipse using the algorithm described by Halir *et al*. [17]. **F**. The points outside either the inner or outer ellipse are considered outliers. The percentage of outliers is displayed on top of the ellipse.

Each movement was recorded for 40 s, resulting in a total data collection time of 5 min 20 s per subject. The recorded EMG data were saved as 2D matrices, the two dimensions representing the 320 electrodes and the recorded time. Successively, we split the data into multiple 100 ms (192 samples) windows with a shift of 31.25 ms (64 samples). The EMG window shift of 64 samples was set because this was the fastest real-time output setting of the EMG recording device (EMG Quattrocento, OT Bioelettronica, Turin, Italy) used in our experiments. The size of the EMG window was chosen as three times the shift. The middle 30% (12 s) of the data for each movement is designated as the test set, while the remaining 70% (28 s) is allocated for the training set. Within the test set for each movement, the first 10% (1.2 s) is further reserved for the validation set. This results in 3 min 44 s training, 1 min 26 s testing, and 9.6 s validation data per participant.

To increase the training data size, we employed three augmentation techniques described in Tsinganos *et al*. [16]:

- Gaussian noise: Gaussian noise was added to the original EMG data with a signal-to-noise ratio of 5 dB.
- EMG magnitude warping: To mimic the effect of movement drifts of the electrode grids, we applied a randomly generated cubic spline to the EMG signals. This introduces non-linear warps to the signal.
- Wavelet decomposition and reconstruction: To further increase the amount of data, we decomposed the EMG signal using wavelets. Before reconstructing the signal, we multiplied the decomposed factors by a constant, ensuring that the reconstructed signal is a novel yet representative (correlated) to the original EMG signal.

The augmentations offered a three-fold increase in training data, resulting in a total per subject of 18 min and 56 s.

### 2.2 Ethical approval

All participants gave their written informed consent to participate in the study. The study was carried out in accordance with the Declaration of Helsinki, except for registration in a database. All procedures and experiments were approved by the ethics committee of the FriedrichAlexander-Universität (applications 22-138-Bm and 21-150B).

### 2.3 Model

Using the EMG data collected, which were synced with the digitally displayed hand kinematics, we trained our previously published model [11], which is capable of accurately and continuously predicting hand kinematics in real-time for each individual participant. The model was implemented in Python using PyTorch (version 2.1.0+cu121) and PyTorch-Lightning (version 2.1.0).

The deep learning model first divided each 100ms EMG window into five individual grids, resulting in five matrices instead of one large one. The model was made out of two parts: a convolutional network and a multi-layer perceptron. We have previously shown [18] that the convolutional network extracts the motor information and projects it into a high-dimensional latent space. The multi-layered perceptron mapped the high-dimensional space into the desired 3D kinematics output.

### 2.4 Latent Space Projection

The latent space of the models (Fig. 2C) are each 2256 dimensional vectors, which, for human comprehension, were projected into only 2 (Fig. 2D). To achieve this, we used the dimensionality reduction technique known as uniform manifold approximation and projection (UMAP) [12]. UMAP was chosen because it is capable of preserving both local and, to some extent, global non-linear structures in the data. This is accomplished by constructing a graph based on similarities among data points, where the similarity is computed using a user-selected metric, such as cosine similarity in our case. The graph is then optimized to reduce its dimensionality while retaining as many local and global relationships as possible. However, the optimization process is inherently stochastic, which can result in different spatial rotations of the data for each subject. To ensure consistent interpretability across subjects, we aligned the 2D embeddings during the optimization process to minimize orientation variations.

The parameters (Fig. 2E & F, Fig. 6B-I, and Fig. 7B) extracted from the manifolds (Fig. 2D) were computed as follows:

- For the temporal behavior analysis (Eq. 1, Fig. 6B & C), the angle (*α*) between an embedding point (*p*) and the centroid (*c*) of the task manifold was computed using the formula:

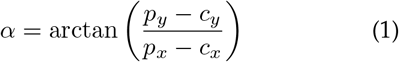

Here, the subscripts *x* and *y* refer to the respective components along the xand y-axes of the points (*p*) and the centroid (*c*). The angles between the centroid and each point on the task manifold are then phase unwrapped. For perfect circularity, this results in a straight line (Fig. 6B), whereas imperfect circularity leads to a discontinuous line. The measure of fitness (*R*^2^) was computed between the phase unwrapped angles and a linear fit for each task and for each subject (Fig. 6C).

- Each manifold (Fig. 2D) was fitted with an ellipse using the algorithm described in Halir *et al*. [17]. To enclose the manifold within an ellipse, the fitted axes (semi-minor and semi-major) were multiplied by a factor of 2 (Fig. 2E, Fig. 6D). To further delineate the anticipated shape of the actual manifold, we used a secondary ellipse measuring 0.5 times the dimensions of the fitted ellipse. This smaller ellipse, in conjunction with the larger one, formed a 2D band-an ellipsoidal annulus that represented our hypothesized manifold shape (Fig. 2E). Using this band structure created by the larger and smaller ellipses, we computed the number of outliers (Fig. 2F). The number of outliers was divided into two groups depending on whether they were encased by the band or outside of it (Fig. 5). To assess how well the manifold adheres to the 2D ellipsoidal band, we computed the closest distance from each point on the manifold to the band. The sum of these squared distances is small when the band is closely followed and increases as the spatial arrangement of the latent space becomes more chaotic (Fig. 7B).
- Sparsity (*s*, Eq. 2, Fig. 6F & G) was defined as

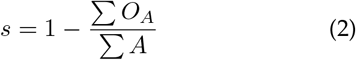

1 minus the division between the sum of all overlap areas (*O*_*A*_) and the sum of the total areas (*A*). This measure yields a value of 1 in cases where there are no overlaps, while a value of 0 indicates that all areas overlap and that tasks cannot be distinguished from each other.

- The number of motor dimensions (Fig. 6H & I) was set to 8 at the start, which corresponds to the total number of tasks. For each overlap found between two tasks, the amount of motor dimensions was reduced by 1, because the merged tasks were still separable from the rest of the tasks.

### 2.5 Conformal Prediction

Conformal prediction [19] is a framework for machine learning that provides calibrated prediction sets that, in turn, can be used as a proxy for uncertainty quantification. We used Regularized Adaptive Prediction Sets (RAPS) [20], as it showed visually reliable prediction regions that adapt well to the underlying data.

The fundamental concept behind RAPS is to construct prediction sets around individual predictions made by a model. These sets are designed to capture the true target value with a certain probability. Unlike point predictions, these prediction sets provide a range of possible values and quantify the uncertainty associated with the model’s predictions. In this study, we used prediction sets with a 95% probability of containing the true class within the measured data. This approach allowed us to quantify whether a prediction is considered certain — indicated by a single prediction within the set — or uncertain, denoting multiple predictions within the set (Fig. 4C & 5B).

### 2.6 Comparison Models

We compared our model with two standard machine learning approaches, a linear discriminant analysis model to separate movement tasks and a ridge regressor to predict proportional movement. Both models were trained with the following features computed for each 100ms EMG window:

- mean absolute value
- root mean square value per electrode
- slope sign change
- waveform length
- zero crossings

This resulted in 324 features for each EMG window. The features were standardized using the statistics obtained from the training set. During testing, the same scaling parameters derived from the training set were applied.

### 2.7 Statistical Measures

The statistical analysis (Fig. 3C, 4D, 6C, 6G, and 6I) was performed using Welch’s t-test because the normal distribution assumption of the data could not be guaranteed. A p-value of below 0.05 was considered significant.

**Fig. 3:**
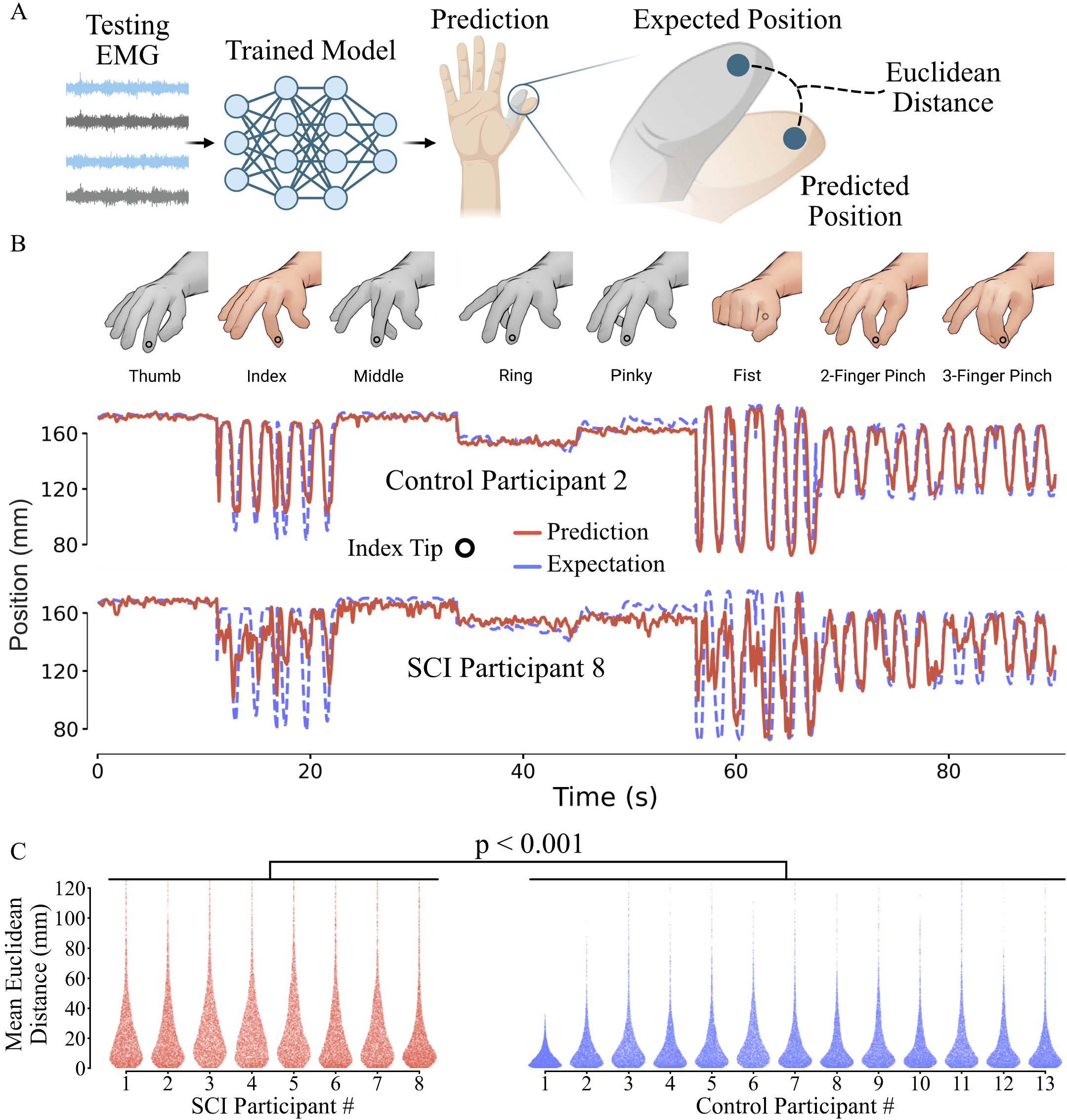
**A**. The predictions derived from the test dataset are compared against the camera recorded kinematics of a repersentative uninjured participant by utilizing the Euclidean distance of the predicted and measured individual fingertip trajectories. **B**. Comparison of predicted and expected index fingertip positions (y-axis) for an uninjured (n. 2) and an SCI afflicted participant (n. 8) in the test set. The colored and grayed-out indications at the top denote whether the index finger is active or not during a task. **C**. Mean Euclidean distance for all subjects in mm. Control group participants show an error of 13.6 ± 15.4 mm, while the participants with SCI demonstrate an error of 23.9 ± 23.2 mm. Statistical significance has been confirmed (p *<* 0.001) using a Welch’s t-test with a total of 120,980 data points with 5,762 per participant (74,884 from control group and 46,096 from SCI group).

**Fig. 4:**
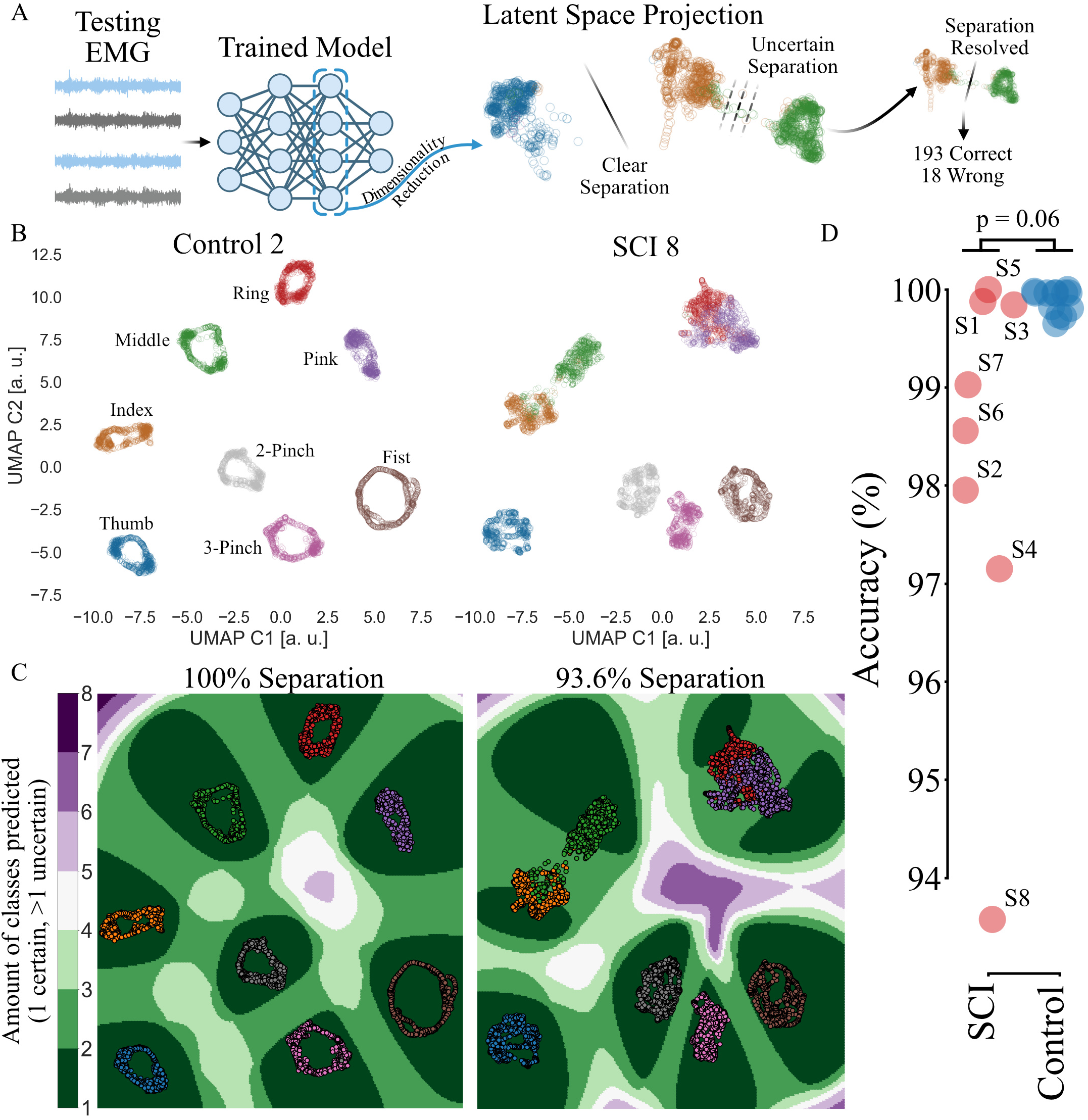
**A**. The latent spaces of the trained networks are projected into 2D using UMAP [12], a non-linear dimensionality reduction technique that preserves local and global information. A support vector classifier is used to separate the clusters in the 2D projection. We observed that for some participants with SCI the tasks could not be entirely separated and appear to be intertwined. To account for this uncertainty in classification, we applied conformal prediction to get prediction sets instead of only one prediction per data point. The prediction sets are 95% probable to contain the correct class inside. If a prediction set is uncertain (i.e., it contains more than one class), we can solve the uncertainty by looking at previous predictions that have been certain and thus making an informed estimate of the class. **B**. Example 2D projections showing the 2nd uninjured (control) and 8th spinal cord injured participant. Datapoints are colored according to each movement. **C**. Visualization of prediction uncertainty for the example latent space projections from B. The colorbar indicates the number of classes within the prediction set. A set is considered certain if it contains only one class. A prediction set with more than one class represents uncertainty in the model. **D**. Accuracy over all participants. Control participants achieved 99.8% while those with a SCI 98.3% accuracy. The points from the SCI group are denoted by an “S” followed by the participant id. No statistical significance (p = 0.06) has been found using Welch’s t-test.

## 3 Results

We collected EMG data from two groups. The control group consisted of 13 uninjured participants while the SCI group consisted of 8 people with motor-complete SCI (C3-C6), classified as AIS grades A to C [2]. Participants were asked to attempt various hand movements displayed on a screen, such as grasping, pinching, and flexing each individual finger while recording their EMG signals from the forearm. Each movement was displayed through a virtual hand. Using the EMG and synchronized hand kinematics data, we trained our previously published AI model [11] capable of real-time kinematics prediction for each participant individually to assess whether proportional control can be used in individuals with SCI and if so for which movements.

### 3.1 Hand Kinematics Prediction

We evaluated the prediction of the kinematics using the Euclidean distance between the expected and estimated hand kinematics (Fig. 3A). Fig. 3B shows the prediction for two representative participants (the 2nd control participant and the 8th participant with SCI). The colored hands, shown at the top, illustrate the active participation of the index finger in the executed movement. For instance, tasks such as grasping and pinching activate the index finger, while tasks such as flexion of the thumb do not. For an assessment of relative predictive capacity, we show the index fingertip prediction versus the expected values throughout the test set, encompassing all executed movements (Fig. 3B). The aggregated Euclidean distance across all subjects is depicted in Fig. 3C. The mean distance for the control participants was 13.6 ± 15.4 mm, while for the SCI participants it was 23.9 ± 23.2 mm. Each subject contributed 5,762 datapoints, resulting in a total of 120,980 for the entire dataset (74,884 from the control group and 46,096 from the SCI group). The means were found to be significantly different (p *<* 0.001, Welch’s t-test).

In the representative example depicted in Fig. 3B, it is evident that the spinal cord injured participant demonstrated proportional control over both pinching and, to some extent, the action of closing and opening the fist. This indicates their ability to generate voluntary and task-modulated signals despite having a motor-complete SCI. Despite some noise, the AI successfully interpreted this activity and generated the intended hand kinematics, underscoring the viability of proportional control in individuals with SCI.

### 3.2 Latent Space Analysis

After examining the prediction of the AI model (that is, the correlation between estimated and expected kinematics), we examined the internal representation learned by the AI from the EMG signals, which corresponds to the latent space of the model (Fig. 4, 5, 6, and 7). Because the latent space of the model contains a human-incomprehensible amount of dimensions, we projected the latent space using UMAP into 2 dimensions. Briefly, this method enables the visualization of high-dimensional nonlinear data into a discernible lowdimensional output (see Methods for details). The UMAP output was analyzed for both the control and SCI groups. We compared the spread of the clusters for each digit that was learned by the AI, their overlaps, and the shape of the continuous prediction relative to time and space. We observe a lack of overlap between the clusters of the control group compared to the SCI group. Participants with SCI showed overlaps between related tasks (e.g., ring and pinky fingers are anatomically close together) (Fig. 4B), which may indicate that at the neural level there is a low number of spared motor units encoding that specific task. In addition, the clusters of the control group are arranged in circular shapes, whereas in the SCI group they generally appear more chaotic, which indicates that sinusoidal movements (e.g., continuous change in index flexion and extension position) are not discernible as well as for uninjured participants.

**Fig. 5:**
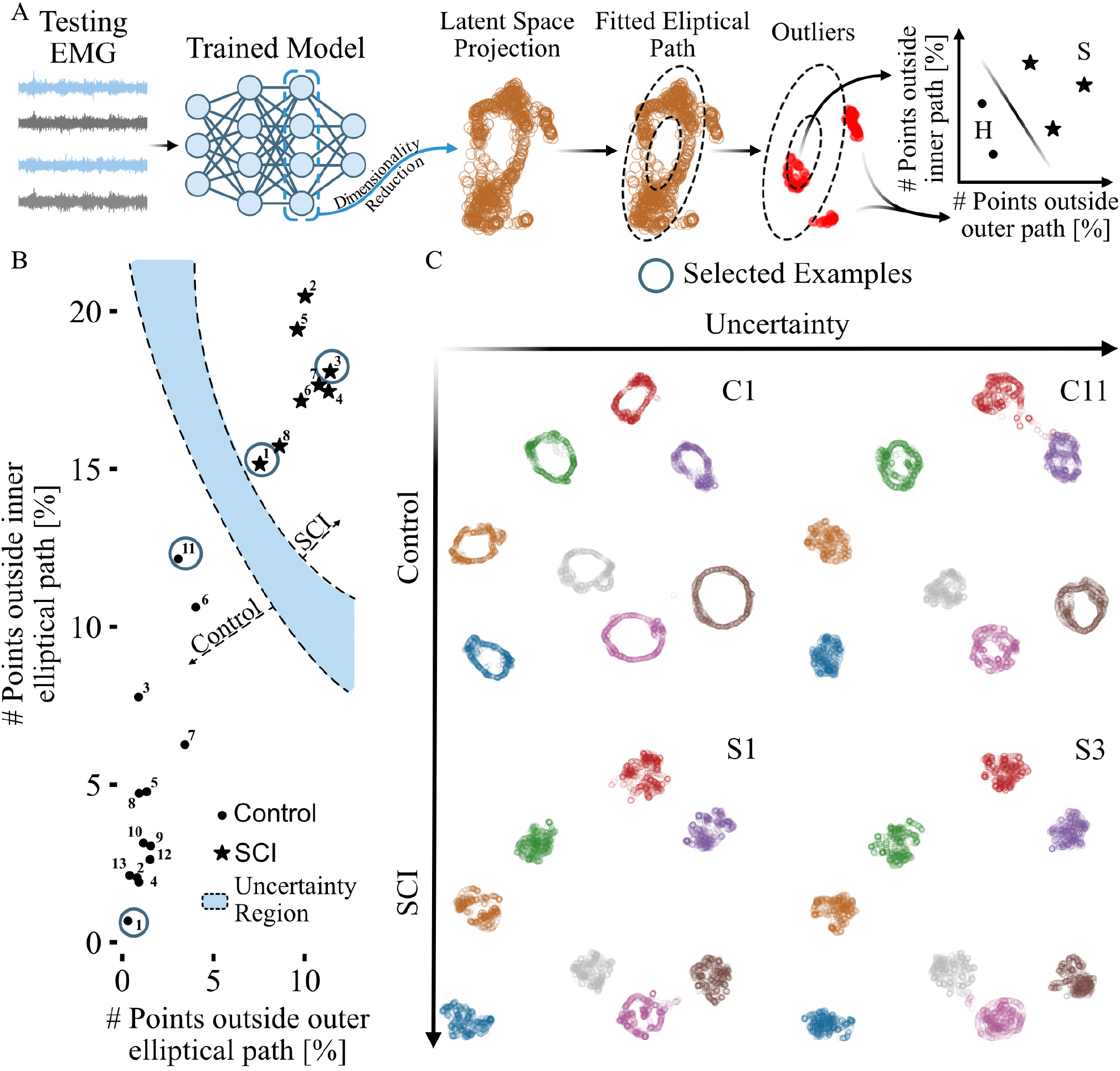
**A**. The high-dimensional latent space was projected into 2D using UMAP [12]. Each task manifold was fitted with a 2D band representing our hypothesized optimal shape (see Methods). The number of outliers outside the band and those enclosed by it was computed and averaged for each subject. **B**. Using the outlier data obtained as described in A, a support vector classifier was trained to separate the participants based on the two features. Uncertainty was gauged using conformal prediction (see Methods). The numbers denote the participant ID. **C**. For interpretability, one certain and one uncertain example is shown for the control and SCI group. The participant IDs are indicated in the upper right corners.

**Fig. 6:**
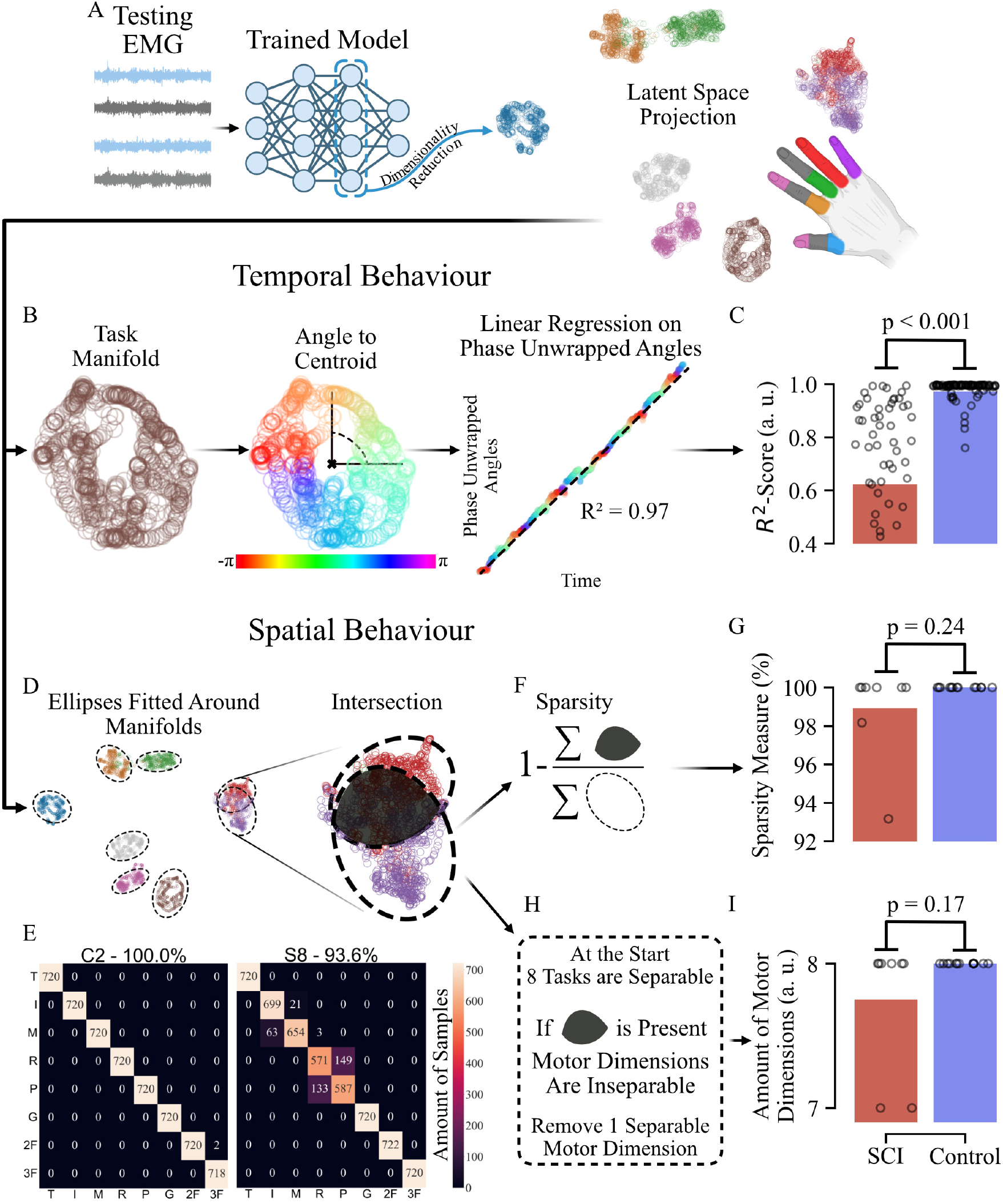
**A**. The 2D projection of the latent space (example shown for the 8th SCI subject) was analyzed for both temporal (B & C) and spatial (D-H) behaviour. **B**. The angle of each point of the manifold of the task relative to the centroid of the manifold was computed using Eq. 1. The resulting angles have then been phase-unwrapped, which, in case of being the result of a circular path, would result in a straight line. This assumption was used to calculate the *R*^2^-Score of the angles in a linear regression fit. **C**. Analysis described in B was performed on all participants. The differences between uninjured participants and those affected by an SCI were found to be significant (p *<* 0.001) using Welch’s t-test. **D**. For the spatial behaviour analysis, we fitted ellipses around each task manifold and computed the intersections between them. **E**. Confusion matrices for the 2nd control and the 8th SCI participants. Movement tasks are denoted by a simplified notation (T - Thumb, I - Index, M - Middle, R - Ring, P - Pinky, G - Grasp, 2F - 2 Finger Pinch, and 3F - 3 Finger Pinch). **F**. Using Eq. 2 we calculated the amount of overlaps divided by the total area of the manifold (see Methods for more details). **G**. Sparsity analysis described in F was carried out on all participants. The differences between uninjured and participants with SCI were not found to be significant (p = 0.24) using Welch’s t-test. **H**. The amount of separable motor dimensions was considered the maximum of tasks executed (8). If there is overlap between two tasks, the amount is reduced by 1. **I**. Motor dimension analysis described in H was carried out for all subjects. Differences between control and SCI participants were not found to be significant (p = 0.17) using Welch’s t-test.

**Fig. 7:**
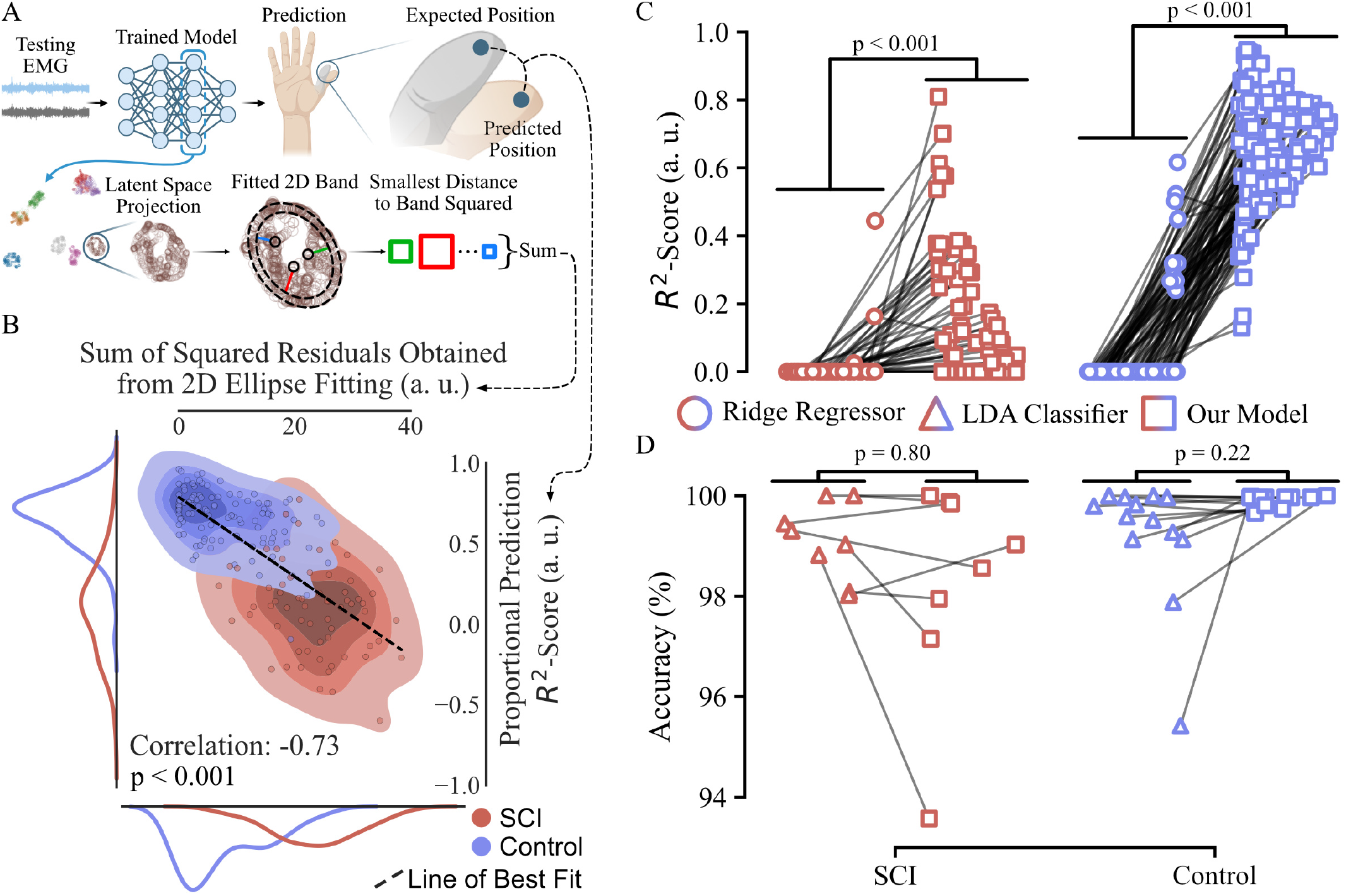
**A**. Correlation between the sum of squared residuals (distance from each point to 2D ellipsoidal band) and the *R*^2^-Scores computed for predicted versus expected fingertip positions across the test set. The y-axis represents the average prediction *R*^2^-Score, while the x-axis shows the sum of squared residuals. **B**. Correlation scatter plot with a superimposed line of best fit, showing the relationship between the two measures. Additionally, the density of the points is shown by a 2D kernel density estimation (KDE). Marginal KDEs are also displayed for each axis. The correlation coefficient was -0.73, indicating a significant negative relationship between the amount of deviation of the latent space from the 2D band (sum of squared residuals) and the prediction of kinematics (p-value *<* 0.001). **C**. Comparison between our model and a ridge regressor model, where each point on the graph represents the *R*^2^-Score of a task’s proportional prediction. The connecting line links the corresponding task scores between the ridge regressor and our model. Statistical analysis revealed significant differences between models for both SCI and control participants, with p-values less than 0.001, as determined by Welch’s t-test. **D**. Comparison between our model and a linear discriminant analysis model (LDA), where each point on the graph represents the accuracy of the movement task separation of a participant. The connecting line links the corresponding scores between the LDA and our model. Statistical analysis revealed no significant differences between the models for both SCI and control participants, with p-values greater than 0.05, determined by Welch’s t-test.

#### 3.2.1 Classification Between Tasks

The 2D projected latent space was clearly separable (Fig. 4) into the different task clusters. The organization of the latent space into sparse task clusters represents a logical and intrinsic initial stage for the AI model during training. Without this step, the model would lack the ability to perform targeted searches for patterns indicative of spatial positions, thereby hindering its capacity for movement-specific analysis. To classify the movements, we used a support vector classifier with conformal prediction to assess the uncertainty level in the separation learned by the network. Conformal prediction (refer to Section 2.5) allows us to discern instances where the model lacks confidence in its separation, thus showing up as overlaps of clusters in the latent space (Fig. 4A). For consistency, we selected the same participants (2nd control and 8th SCI) to illustrate the projections of the 2D latent space both before (Fig. 4B) and after applying conformal prediction (Fig. 4C). A prediction set containing more than one class was considered uncertain, while a set comprising only one prediction was deemed certain. The aggregated separation accuracy is presented in Fig. 4D. The uninjured participants had a mean separation accuracy of 99.8%, while for those with an SCI it was 98.3%. No significant differences were observed between the uninjured and the spinal cord injured participants (p = 0.06, Welch’s t-test).

#### 3.2.2 Classification Between Control and SCI

Latent space projection could be further used to distinguish between uninjured and spinal cord injured participants with 100% accuracy (Fig. 5) despite the networks not receiving any input data regarding the groups (e.g., if a specific participant suffered a SCI or not). For each task cluster, we computed a 2D idealized path in the shape of a band (Section 2.4, Fig. 5A). Outliers situated both outside and within the 2D path were used to determine the classification of participants as either suffering from a SCI or not (Fig. 5A). A support vector classifier was used in conjunction with conformal prediction (Section 2.5) to separate SCI from control participants based only on the amounts of internal and external outliers from each task clusters (Fig. 5B). To facilitate the understanding about the uncertain regions, which correspond to movements that were not easily separated by the AI model, we provide latent spaces for “highly certain” and “highly uncertain” participants from both the control and SCI groups in Fig. 5C.

Although the network has not been trained to distinguish between participants with and without SCI, the latent space effectively encodes this information from its shape alone. This is not surprising considering that the amount of spared MU activity in SCI is significantly smaller than in healthy [5].

#### 3.2.3 Manifold Parameters

We first examined the temporal behaviour of the task manifolds (Fig. 6A & B). We wanted to understand whether the appearing circular shapes have been constructed randomly or if there is an underlying rotational path due to physiological dynamics present in the EMG signals. To test this, we computed the phase unwrapped angles of each point forming the circular shape and calculated the coefficient of determination (*R*^2^-Score) to see how similar the result is to a straight fitted line (see Section 2.4 for details). The averaged results for both participant groups are displayed in Fig. 6C) and have been found to be statistically significant (p *<* 0.001, Welch’s t-test).

The other two parameters extracted are concerned with the spatial behaviour of the latent spaces. For each task manifold, we computed an ellipse that best fitted the latent space projection (Section 2.4, Fig. 6D) and calculated the intersection area between the ellipses. The separation accuracy for the 2 representative subjects (2nd control 100% and 8th SCI 93.6%) can be seen in Fig. 6E. Using the intersections and the overall area of the ellipse, we computed the sparsity measure (Fig. 6F) which we displayed averaged over all participants in Fig. 6G. The difference in separation accuracy between the control and SCI group was not found to be statistically significant (*p* = 0.24, Welch’s t-test).

Given the presence of overlaps, we computed the number of separable motor dimensions for each participant by initially assuming that each participant has as many motor dimensions as the number of tasks performed (total of 8 tasks) and subtracting 1 each time an overlap was detected (Fig. 6H). The average results for both groups are displayed in Fig. 6I and are not statistically significant (*p* = 0.17, Welch’s t-test).

This demonstrates that the network has successfully learned to distinguish individual movement tasks from each other for the entire control group and the majority of participants with SCI.

### 3.3 Correlation Between Prediction and Temporal Behaviour

Based on the prevalence of a predominant circular arrangement of the movement task manifolds in the control group compared to the SCI group (Fig. 6C), we evaluated the correlation between the amount of deviation (sum of squared residuals) of the latent space from a 2D fitted ellipsoidal band and the prediction accuracy of the hand kinematics (Fig. 7A & B). We computed the average *R*^2^-Score between the predicted and expected fingertip positions for all movements in the test set and correlated it against the sum of squared residuals (Fig. 7A). In Fig. 7B, we display the correlation scatter plot with the best fit line superimposed on it. For a better understanding of the density of the points, we also computed the kernel density estimation (KDE) and displayed both the 2D KDE and the marginal KDE for each respective axis. The correlation achieved was -0.73 (p *<* 0.001).

We note that movements that do not follow a circular path (the sum of squared residuals is high) exhibit a negative correlation with their proportional controllability. This correlation suggests the potential utility of employing such analysis for the detection of proportionally recoverable movements which can be used for targeted neurorehabilitation.

### 3.4 Comparison With Standard Models

The last test performed was to compare our model with two standard models. A ridge regressor to proportionally predict all hand joints (Fig. 5C) and a linear discriminant analysis model to classify individual hand movements from each other (Fig. 5D). For regression, we present the mean *R*^2^-Score for each movement task, which yields a total of 96 data points for the control participants and 64 data points for participants with SCI. In the case of classification, we report accuracy for each participant, resulting in 13 and 8 data points for control and spinal cord injured participants respectively.

Our model demonstrates significantly better regression performance (p *<* 0.001, Welch’s t-test), while showing no significant difference from linear discriminant analysis in classification (p = 0.80 or 0.22, Welch’s t-test).

## 4 Discussion

We analyzed the feasibility of decoding proportional control of hand movements for individuals with a chronic motor-complete SCI, using a convolutional neural network. Our system only requires EMG data recorded while attempting different hand movements. The output of our AI model can provide insight into which motor dimensions (hand movements) are spared and proportionally controllable after SCI. The results from the SCI group were compared with a cohort of uninjured control participants.

We found that the prediction of hand kinematics for certain movements attempted by individuals with SCI was similar to that of uninjured participants (Fig. 3). Moreover, by analyzing the latent space of the prediction models, we observed that such movements describe distinctive circular manifolds. We show that the deviation of a task manifold from an ellipsoidal path is negatively correlated (-0.73, Fig. 7B) with the accuracy of the prediction. Our system can thus produce a measure of how well a movement can be proportionally predicted (Fig. 6 & 7).

### 4.1 Circular Latent Space Projections Correlate with Proportional Prediction

Given that all participants in the SCI group have been unable to use their hands for at least several years, we anticipated that the error (mean Euclidean distance) between the estimated and expected hand kinematics would be statistically higher for them compared to the control group (Fig. 3). The network’s ability to proportionally decode some movements in the SCI group (Fig. 3) is consistent with our previous findings, where we demonstrated similar capabilities in a smaller subset of movements [4], [5].

Although not all movements attempted by participants with SCI could be proportionally decoded, we found that the majority of movement tasks were distinguishable from each other using the latent space of the neural network (Fig. 4 and Fig. 6G & I). The separation accuracy remained consistent between the uninjured individuals and those with SCI, with no significant statistical differences observed between the two groups (Fig. 6G & I). Our findings suggest that participants with motor-complete chronic SCI were able to produce signals varied enough to differentiate between multiple movements. Our previous study [5], in which we performed a real-time decomposition of sEMG signals during movements analogous to those examined in this work, found that most tasks shared a limited number of MUs. The sparsity findings (Fig. 6G & I) in this study also support this notion. It is easier to separate signals with less overlap (fewer MUs) than those with a higher level of overlap, as expected from uninjured participants.

Despite not training the hand kinematics decoder models to distinguish between individuals with and without SCI, we demonstrate that the latent space encodes this information with 100% accuracy (Fig. 5). Not only do we find the latent space to be structured, but we could also extract different physiological parameters from it. We showed that individual movement manifolds are temporally arranged in a circular path, reflecting the cyclical nature of attempted movements (Fig. 6B & C). Furthermore, we observed that the separation of tasks is sparse, with no significant differences between the two groups (Fig. 6D-I).

A circle, however, is characterized not only by having its constituting points follow a temporally circular path, but also by the points being spatially equidistant from the center. We demonstrate that the deviation from an elliptical band fitted to an individual movement task manifold is negatively correlated (-0.73) with the accuracy of movement proportional decoding. This observation implies that as the chaotic nature of a manifold increases, there is a corresponding rise in the error rate of its proportional prediction (Fig. 7A & B).

### 4.2 Necessity of Deep Learning

We recorded the forearm muscles of participants with and without SCI using 320 electrodes. Our previous research has shown that decoding hand movements is enhanced with a larger number of electrodes [21], as it eliminates the need to precisely define the recording area, which is particularly advantageous when dealing with the territories of unknown spared MUs in participants with SCI. Having this large amount of data warrants the question of the necessity of a deep learning model, as a machine learning variant might be just as effective and easier to implement in practice.

Indeed, we observed that our deep learning network performs similarly to its machine learning counterpart in movement classification (Fig. 7D). However, the machine learning model struggles to proportionally predict movements in both the injured and uninjured groups, a task at which our deep learning network excels (Fig. 7C).

The success of the algorithms utilized for classifying various hand movements (Fig. 7D) implies that they autonomously acquired the ability to filter out non-movement relevant MU signals while retaining those encoding motor intent. This notion is further substantiated by the sparsity measure (Fig.6G) and the quantification of separable motor dimensions (Fig.6I).

### 4.3 Potential Clinical Application

Due to a negative correlation of -0.73 (p *<* 0.001, Fig. 7B) observed between the deviation from circularity of the latent space embeddings and the accuracy of the proportional prediction, we propose that our system could serve as a test to identify potential movements that can be fully restored through an assistive system and direct monitoring of the spared neural pathways during rehabilitation.

The proposed test would involve patients trying to replicate the movements detailed in this study, while a model predicts their kinematics based on recorded sEMG signals. Subsequently, the UMAP projection of the latent space could help identify circular manifolds. This identification process could be performed by medical professionals or by using algorithms, as described in Section 2.4. Implementing such a test could help medical professionals in planning and monitoring rehabilitation plans that specifically target movements with the potential for proportional control by the patient.

Another significant clinical application is that even when a movement is not proportionally controllable, our system demonstrates reliable classification accuracy (*>* 93%, Fig. 4). Consequently, it can be at best used to proportionally decode a movement and at worst classify it as being different from the other movements learned. This can be used as the control input to an assistive device that has substantial potential to enable patients to be independent in daily living tasks and significantly reduce dependence on caregivers.

However, most importantly, our results emphasize the need to assess whether a patient who cannot move without assistance can produce volitional motor signals that are detectable by EMG. This aspect is currently not part of the standard SCI assessment procedure [2], despite the potential presence of proportionally controllable EMG signals postinjury. Such signals can inform assistive devices, such as an exoskeleton or orthoses, on which movement to generate. This distinction would not only significantly impact the psychological well-being of patients by informing them that they can regain movement with an assistive device, but it would also benefit physicians. Doctors can design and plan more effective rehabilitation treatments that target specific movements that the patient can control. Furthermore, this distinction would benefit researchers by eliminating the time-consuming examination of each potential study candidate that would be necessary without such an assessment. Therefore, we suggest that the motor-complete diagnosis of SCI also assesses the ability to generate EMG signals. If a patient can produce intentional EMG signals, they are considered EMG incomplete; otherwise, they are classified as EMG complete. The need for a more refined classification of patients with motor-complete SCI has also been proposed by Sangari *et al*. [22], who found a correlation between the presence of spasticity and subsequent improvement in the assigned grade of injury for the patient. Spasticity, indicative of spared MUs post-injury, can be evaluated through noninvasive methods such as EMG.

## 5 Conclusion

Our system demonstrated the ability to predict certain movements proportionally in participants suffering from a motor-complete chronic SCI (Fig. 3) and achieved a classification accuracy above 93% for the 8 attempted movements (Fig. 4). Analysis of the latent space using UMAP revealed that the internal representation learned by the model can perfectly differentiate between participants with or without SCI (Fig. 5). Furthermore, examination of the latent space indicated that movement encoding occurs in sparse manifolds, displaying a circular pattern specifically when movement could be decoded proportionally (Fig. 6 & 7).

Our proposed model holds promise as a motor intent decoder for individuals with motor-complete SCI. It consistently classifies described motions and, in some cases, even provides a proportional regression signal that can be used for an assistive device.

Moreover, considering the correlation between the circular path of the latent space task manifold and the accuracy in proportional control, we suggest that this correlation could serve as a test to identify motor dimensions amenable to proportional control. Implementing such a test could help medical professionals develop more targeted and customized rehabilitation plans to better assist patients.

## Data Availability

All data produced in the present study are available upon reasonable request to the authors

## Acknowledgments

This work was supported by the Bavarian Ministry of Economic Affairs, Regional Development and Energy (StMWi) under grant MV-2303-0006, by the European Research Council (ERC) through the project GRASPAGAIN under grant 101118089, and by the German Federal Ministry of Education and Research (BMBF) through the project MYOREHAB under grant 01DN23002 to ADV.

The authors further gratefully acknowledge the scientific support and HPC resources provided by the Erlangen National High Performance Computing Center (NHR@FAU) of the Friedrich-Alexander-Universität Erlangen-Nü rnberg (FAU). The hardware is funded by the German Research Foundation (DFG).

